# First-born twin has a higher risk of acute leukemia in a population-based assessment of cancer in twins in California, and lower than anticipated rate of twin concordance

**DOI:** 10.1101/2024.07.11.24310290

**Authors:** Eric M Nickels, Naying Zhou, Joseph L Wiemels

## Abstract

We assessed cancer concordance, cancer incidence in the healthy twin of cancer probands, and cancer risk in relation to birth order in pediatric and adolescent/young adult twins via a population-based study in California from 1982-2022. Twin subjects born in California between 1982-2017 who were diagnosed with leukemia from 0-39 years of age were identified through linked birth and California Cancer Registry (1988-2022) data. Two concordant-twin leukemias cases were identified across 255 total twin pairs with leukemia for an overall rate of leukemia concordance of 0.9%. One concordant twin pair was identified among 199 pairs with lymphoid leukemia (0.5%) and one within 34 pairs with acute myeloid leukemia (2.9%). A significant association was identified between twin plurality birth order and development of leukemia (OR 1.18, 95% CI 1-1.39, P=0.049), an effect which was strongest in lymphoid leukemias (2.21, 1.44-3.39, P=1.65e-4). Assessment of DNA methylation markers associated with birth order showed significantly reduced methylation in first-born twin cases compared to second-born (P=8.53e-12) in a subset of 41 twins discordant for lymphoid leukemia. Overall cancer concordance in twins was comparable to the lower range of previous estimates from different world regions. Concordance in lymphoid leukemias was quite lower than expected, indicating concordant leukemia is rarer than previously appreciated. We identified a strong association between twin plurality birth order and development of pediatric cancer. While the underlying cause of this finding is uncertain, we identified significant differences in DNA methylation at previously described sites associated with birth order, suggesting a similar biological mechanism.

## INTRODUCTION

The study of twins has aided in estimations of the germline and somatic genetic contribution to many childhood cancers^1^. In the case of pediatric leukemia, twin studies helped to identify a unique disease etiology in which the initiating somatic genetic translocation event in leukemia development occurs in the intrauterine period and subsequently passes between twin siblings through shared placental circulation^2-5^. This underlying disease mechanism in part explains the relatively high reports of twin concordance for pediatric leukemias, which ranged from 2.5%^6^ to as high as 25%^7^. Given these ranges, and the potential for bias from subject selection, twin concordance in acute lymphoblastic leukemia (ALL) and acute myeloid leukemia (AML) have been estimated at approximately 10%^8^. Importantly, the presence of these early genetic translocations is not sufficient for leukemia to develop^9^, indicating additional genetic, epigenetic and environmental factors are necessary for malignant transformation.

Childhood cancers are thought to arise from both inborn genetic and environmental causes during development. Familial aggregation studies suggest that shared inherited predispositions to cancer and shared environment can play a role, but it can be challenging to differentiate these roles. Elevated disease concordance in monozygotic (genetically identical) compared to dizygotic twin pairs suggests a stronger contribution of *genetic* influence on cancer development. In this manner, these studies can play an important role in understanding childhood cancer etiology and risk factors.

Given the rarity of most types of childhood leukemia, however, estimation of twin concordance has remained challenging. Even in the case of infantile ALL, which is reported to have a twin concordance rate of 100%^8^, rare case reports argue this number may be lower than previously appreciated^10^. While selected large cohorts allow for greater enrollment of twin subjects with leukemia, these studies are at risk of inaccuracies resulting from ascertainment bias^6^. In contrast, population-based studies allow for the assembly of large cohorts of individuals in an unbiased fashion. However, population-based approaches are limited by the associated costs and resources required to assemble sufficient sample sizes of rare disorders with sufficiently detailed diagnostic information. The Nordic Twin Study^11^ is an example of a population-based approach utilizing twins to investigate familiar risk of cancer, providing reports on twin concordance rates for a diversity of cancer types including hematologic malignancies with an estimated heritability of the liability to develop hematologic malignancy of 24%^12^.

These population-based twin studies have focused on the relatively genetically uniform Scandinavian population. Investigation of twin concordance in a genetically diverse population would further aid in the identification of a more broadly applicable estimation of inherited contributors to childhood leukemia development. Here, we seek to Identify pediatric and adolescent/young adult (AYA) twin leukemia concordance rates through a population-based registry assessment of the State of California from 1982-2022.

## MATERIALS AND METHODS

### Identification of twin pairs

This study was approved by Institutional Review Boards at the California Health and Human Services Agency and the University of Southern California. Data analysis was conducted using R (version 4.2.3)^13^. We investigated combined registry data including birth records from the Birth Master Statistical File (BMSF) from California from 1982 to 2017 and cancer diagnosis records from the California Cancer Registry (CCR) from 1988 to 2022 to identify twins in which at least one sibling was diagnosed with pediatric/AYA leukemia. Subjects were linked between registries through a unique family identifier. Twins were identified as siblings within a family unit sharing the same birth date. Higher order multiples were excluded from analysis.

### Characterization of cancer diagnoses

Included age of cancer diagnoses ranged from 0-39 years. Diagnoses were categorized by ICD-O-3 histology codes into 12 broad groups and 69 recode subgroups following the International Classification of Childhood Cancer, Third Edition (ICCC-3, November 2012; Table S1). The leukemia group included five recode subgroups that were included in analysis. Concordant cases were defined as both siblings in a twin unit having a diagnosis in the same recode subgroup, whereas discordant cases were those in which siblings did not share a recode subgroup diagnosis. Cross-cancer concordance was defined as both siblings within a twin pair having diagnoses in separate recode categories.

### Statistical Analysis and determination of concordance rates

Concordance rates were defined as the total number of concordant twin cases divided by the sum of concordant and discordant twin cases. Concordance rate across all cancer types (including non-leukemia diagnoses) are show in table S2. To calculate concordance for all diagnoses, twin units were only considered once, even if an individual had multiple diagnoses in separate broad cancer subgroups or recode subgroups. For subgroup specific analysis, individuals with multiple cancers were assessed separately for each diagnosis based in corresponding broad and recode subgroups. Standardized incidence ratios (SIRs) were calculated based on a previously published method^14,15^.

Fisher’s exact test was used to investigate the association between birth plurality order (i.e., the birth order within a twin pair) and cancer development (Table S2). A paired student’s T-test was used to assess associations between birth weight and cancer development by all cancers and by broad subgroups. Standardized incidence ratios (SIRs) were calculated based on a previously published method^14,15^. Further details on methods can be found in the Supplemental Methods.

Zygosity status was not available in the registry data. The percentage of monozygous twins was estimated based on the assumption that dizygotic twins are equally likely to be represented as same sex or opposite sex twins^6,16^. Thus, we estimated the number of monozygotic twin pairs as the number of same sex pairs minus the number of opposite sex pairs, and dizygotic pairs as double the number of opposite sex pairs as follows:

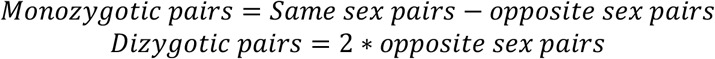

For the purposes of concordance calculations, we assumed all same-sex concordant twin pairs were monozygotic.

## RESULTS

### Cohort description

The combined BMSF and CCR data included 50,982 family units with a total of 238,860 individuals. There were 53,283 individuals with cancer diagnoses and 83,426 unaffected siblings, along with 51,151 mothers and 51,000 fathers. There was a total of 1,213 twins identified with at least one member of the twin pair having a cancer diagnosis (Table S3). Of these, there were 255 twin units in which at least one sibling in the pair had a diagnosis of leukemia (Table 1). There were 166 same sex twin pairs, and 89 opposite sex pairs. Of same sex twin pairs, there was a significantly higher number of males (101, 60.8%) than female (65, 39.2%, P = 0.006). The median gestational age for discordant twin pairs was 260 days (range 183-309). The median maternal age was 31 years (range 16-49) and paternal age 33 years (18-60). The majority of twins were born via cesarian section (183, 71.8%) compared to vaginal deliveries (72, 28.2%). There was a total of 265 distinct cancer diagnoses identified across the twin leukemia cohort, 258 of which were categorized as leukemia. This included a total of 200 diagnoses of lymphoid leukemia, 35 diagnoses of AML, 11 diagnoses of chronic myeloproliferative diseases, 8 diagnoses of myelodysplastic syndrome and other myeloproliferative diseases, and 4 diagnoses of unspecified and other specified leukemias. The median age of diagnosis was 6 years (IQR 10, range 0-30). There was no significant association between birth weight and development of leukemia (Table S2).

**Table 1:**
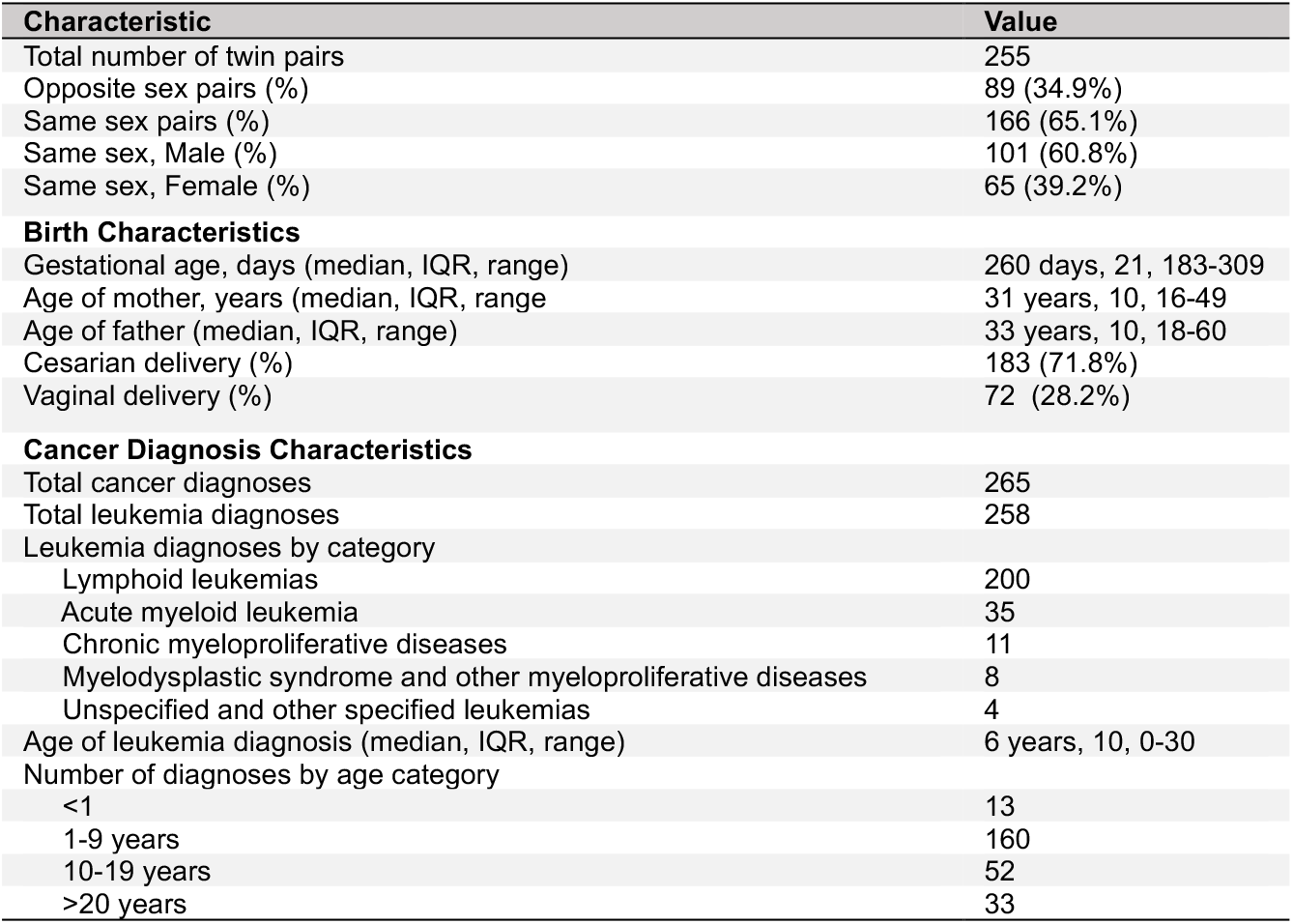
Twin leukemia cohort characteristics.

### Birth Order

We next assessed associations between twin plurality order (being born first or second within the twin pair) and development of leukemia. Across the full twin cohort, there was a significant increased risk of leukemia development associated with being the first-born twin (OR 1.73, 95% CI 1.20-2.52, P =0.003, Figure 1A, Table S4). Within leukemia recode subgroup, lymphoblastic leukemias were most highly associated with being first-born (OR 2.21, 1.44-3.39, P=0.000165), while acute myeloid leukemia showed decreased risk, however this was not statistically significant (OR 0.41, 0.13-1.24, P=0.127, Figure 1) (Table S4). There was no significant relationship between birth plurality order and development of the three remaining recode subgroups.

**Figure 1:**
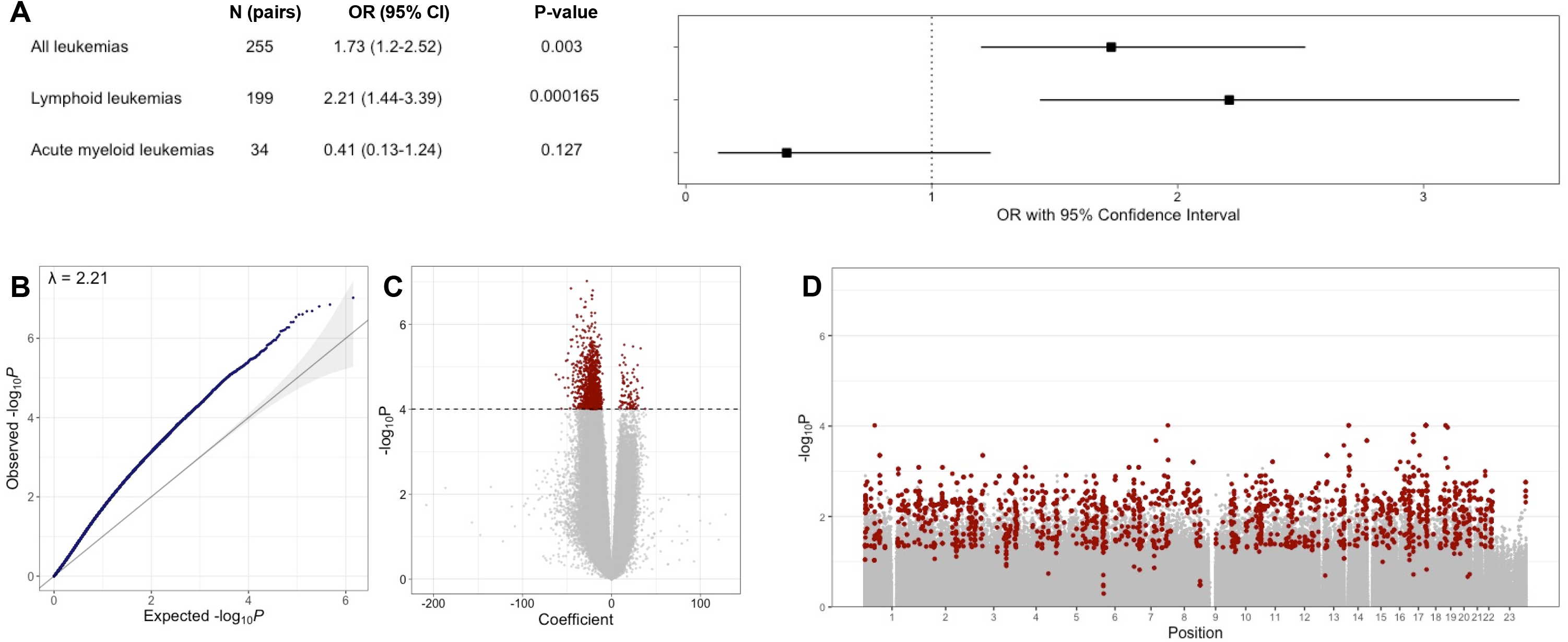
Birth plurality order is significantly associated with development of leukemia in leukemia-discordant twin pairs. (A) Forest plot demonstrating odds ratio from Fisher’s exact test for the association between twin plurality order (first or second born of a set of twin siblings) and pediatric leukemia in leukemia-discordant twin pairs. A significantly higher odds of developing leukemia of any type is seen in first born twins. By disease group, twins discordant for the development of lymphoid leukemia had significantly higher odds of being first born. Twins discordant for development of AML, however, had lower odds of being first born, however this difference was not statistically significant. (B) Quantile-quantile plot of linear regression model for the relationship between DNA methylation status and twin plurality order. Lambda value represents genomic inflation. (C) Volcano plot from the linear regression model showing significance (-log_10_P) by regression coefficient. The 1,394 significant differentially methylated probes (FDR<0.05) are shown in red. (D) Volcano plot showing distribution of -log_10_P values by genomic position. Probes found within the 534 significant differentially methylated regions are highlighted in red.

### Concordant twin cancer cases

Two leukemia-concordant twin pairs were identified in the recode subgroups for lymphoid leukemia and AML (Table 2). In the lymphoid leukemia concordant pair, both siblings were of the same sex and diagnosed at <1 year of age. The AML concordant pair were of the same sex and aged 1-5 at diagnosis. Cross-cancer concordance was identified in a twin pair of the same sex with one sibling diagnosed with lymphoid leukemia at age 1-5 and the second sibling diagnosed with rhabdomyosarcoma at age 6-9.

**Table 2:**
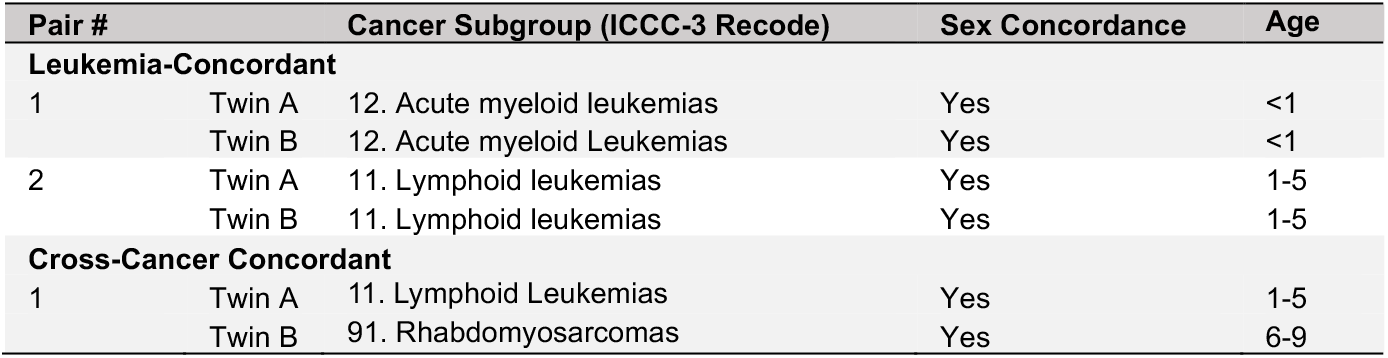
Twin pairs with concordant and cross-cancer concordant leukemia diagnoses. Concordant cases were defined as twin siblings sharing leukemia diagnoses in the same ICCC-3 recode subgroup. Cross-cancer concordant cases were defined as twin siblings with cancer diagnoses in separate recode subgroups, with at least one having a leukemia diagnosis.

The twin concordance rate for all leukemia was 0.8% (2 of 255 twin pairs). For same sex pairs, this rate was 1.2% (2 of 166), and for opposite sex 0% (0 of 89) (Table 3). By recode subgroup, the rate of twin concordance for lymphoid leukemias was 0.5% overall (1 of 199), 0.7% (1 of 134) for same sex and 0% (0 of 65) for opposite sex pairs. For AML, the overall rate was 2.9% (1 of 34), and 5.0% (1 of 20) for same sex and 0% (0 of 14) for opposite sex pairs. Concordance rates were 0% in the categories of chronic myeloproliferative diseases (11 total pairs), myelodysplastic syndrome and myeloproliferative diseases (8 pairs), and unspecified and other specified leukemias (4 pairs). By age groups, the concordance rate for all leukemias for diagnoses under 1 year of age was 8.3% (1 of 12) and 0.63% (1 of 159) for diagnoses age 1-9 years. For age 10 and greater, the concordance rate was 0% (0 of 84). Rates by estimated zygosity status are shown in Table S5. For all leukemias, the estimated monozygotic concordance rate was 2.6%, with rates for lymphoid leukemias 1.4% and AML 16.7%.

**Table 3:** Rates of concordance in twin cancer cases by ICCC-3 broad groups and subgroups. Each tumor is considered separately within each subgroup, and thus subjects with multiple tumors could be considered in independently in separate subgroups. While data is shown only for groups and subgroups with instances of cancer-concordance, the total row includes data from all 12 broad subgroups. Con=concordant, Dis=discordant.

| Category | Same Sex |  |  | Opposite Sex |  |  | All |  |  |
| --- | --- | --- | --- | --- | --- | --- | --- | --- | --- |
|  | Concordant | Discordant | % | Concordant | Discordant | % | Concordant | Discordant | % |
| **All Leukemias** | **2** | **164** | **1.2** | **0** | **89** | **0.0** | **2** | **253** | **0.8** |
| *Lymphoid Leukemias* | 1 | 133 | 0.7 | 0 | 65 | 0.0 | 1 | 198 | 0.5 |
| *Acute Myeloid Leukemias* | 1 | 19 | 5.0 | 0 | 14 | 0.0 | 1 | 33 | 2.9 |
| *Chronic myeloproliferative diseases* | 0 | 8 | 0 | 0 | 3 | 0 | 0 | 11 | 0 |
| *Myelodysplastic syndrome and myeloproliferative diseases* | 0 | 3 | 0 | 0 | 5 | 0 | 0 | 8 | 0 |
| *Unspecified and other specified leukemias* | 0 | 1 | 0 | 0 | 3 | 0 | 0 | 4 | 0 |

### Standardized Incidence Ratios (SIRs)

SIRs were calculated to characterize the relative risk of cancer development in twin siblings of probands diagnosed with a leukemia diagnosis. The SIR for developing a leukemia of the same recode subgroup as the proband in twin siblings was 15.6 (95% CI 1.9-56.4, Table S6). The SIR for development of any cancer in twin siblings of probands with a leukemia diagnosis was 2.9 (95% CI 0.4-10.5, Table S7).

### Birth order DNA methylation analysis

We investigated 41 pediatric ALL-discordant twin pairs who previously underwent genome-wide DNA methylation profiling at birth^17^ for differences in DNA methylation based on twin plurality order. Deconvolution analysis showed no significant differences in nucleated cell proportions between first and second born twins (Table S8, Figure S1). To assess for the relationship between epigenome-wide DNA methylation status at birth and twin plurality order, we used a linear regression model controlling for ALL cases status, birthweight, sex, twin pair number, nucleated cell proportions and array batch effect. Of the 710,010 CpGs assessed, 1,394 significant differentially methylated probes (DMPs, FDR <0.05), reflected by a moderately elevated genomic inflation value of 2.21 (Figure 1B-C), with the top 20 DMPs shown in Table S9. Results from linear regression analysis for all significant results are available in Supplemental Data File 1. There was a total of 534 significant differentially methylated regions (DMRs, Comb-P Sidak P-value <0.05, Figure 1D), the top 20 of which are shown in Table S10. Results from Comb-P analysis are available in Supplemental Data File 2.

We then evaluated CpGs with known association to birth order^18^. Of the 341 birth-order CpGs, 305 overlapped with the 710,010 probes from the EPIC array for the ALL-discordant twin cohort. We next assessed absolute difference between first and second born twins for each CpG (delta-beta values). Mean delta-beta values were significantly lower (hypomethylated) across the 24 twin pairs in which the ALL case was first-born compared to the 17 in which the case was second born (Wilcoxon rank-sum, P=8.53e-12, Figure S2). In stratified analysis splitting those where the cancer proband was born first and those second, first born ALL twins were significantly hypomethylated at these CpG loci, and first-born healthy children were significantly hypermethylated (One-sided Wilcoxon test P=5.1e-8, and 5.0e-6, respectively)

## DISCUSSION

We identified an overall concordance rate of 0.8% for twins with leukemia, which is notably lower than the reported range of 5-25%^8^. By specific diagnosis, we found concordance rates of 0.5% of twins with lymphoid leukemias and 3.1% of twins with AML. While we are unable to confirm zygosity status through the registry data, our estimation of concordance in monozygotic twins for lymphoid leukemias remains low at 1.4%, while that of AML is within the previously reported range at 16.7%. Importantly, this estimation assumes all same sex twin cases concordant for cancer are likely to be monozygotic, and thus is more likely to underestimate monozygotic twin concordance. These results argue that concordant cases of twin acute leukemia are rarer than previously appreciated, which may result from biased recruitment in previously reported cohorts and potential oversampling of rare concordant twins relative to the true population background rate^8^. Even within cases of leukemia diagnosed at less than 1 year of age, which are reported to have concordance rates near 100%, we found a rate of only 8.3%. It is also notable that this study examined twins in a multiethnic birth population in which Latino births represent the majority population; most prior studies being performed in non-Latino whites. The low rate of twin concordance indicates that *acquired* tumor genetic, epigenetic and discordant responses to environmental exposures may contribute more heavily to pediatric and AYA cancer etiology than previously appreciated in pediatric acute leukemias, or at least among those in recent decades in California. In comparison, we identified a concordance rate of 1.1% for all cancer diagnoses, which falls at the lower range of previously reported general twin cancer concordance rates^6^.

Similarly, we identified a counter-twin leukemia specific SIR of 15.6, which is notably below previously published rates. An investigation of 211 twins in the Childhood Cancer Survivor Study^1^ reported a counter-twin SIR of 38.4 (17.3–85.6)^1^, while other studies have reported SIRs as high as 208.28 (54.05-641.09)^19^. These findings would presumably argue leukemia etiology has greater complexity in the interplay between germline genetics and environmental exposures/developmental trajectories than previously appreciated.

Twin studies helped to support the concept of an intrauterine etiology of at least some subtypes of pediatric acute leukemia, in which an initiating mutation occurs in one sibling and is transferred to the second through shared fetal circulation^20^. Molecular studies have since confirmed the presence of identical driver mutations in concordant twin siblings for pediatric ALL of *ETV6-RUNX1*^*3*^, *BCR-ABL1*^*21*^, *TCF3-ZNF384*^*22*^, and high hyperdiploidy^23^ subtypes, along with AML with *KMT2A* rearrangement^24^. The initiating molecular hit is thus shared between twins at birth, with subsequent mutations leading to overt development of acute leukemia with latency as long as 14 years following the initiating event^25^. Here, we identified a pair of same-sex twin siblings concordant for AML at age 1-5 years and a pair of same-sex twin siblings with lymphoid leukemia at less than 1 year of age, which is consistent with this scenario.

In addition, we uncovered a unique association between twin plurality order and leukemia development which has not previously been described. First-born twins were more likely to develop any leukemia compared to second born twins, a finding which was most prominent in twins with ALL. We conducted linear regression modeling to investigate how this relationship might be reflected in DNA methylation at birth in twins discordant, in which we found evidence of genomic inflation indicative of higher-than-expected P-values across all CpG probes on the EPIC array. This would suggest the underlying biological mechanism responsible for the difference in leukemia risk between first and second born twins is reflected in a broad alteration of DNA methylation across the epigenome. The general direction of this alteration is reduced DNA methylation in the first-born twin.

Interestingly, there are prior known associations with birth order and the risk of developing acute leukemia. First born individuals are at increased risk of ALL^26^, while later born (4^th^ or more) individuals are at greater risk of development pediatric AML^27^. In the case of ALL, this inverse association with birth order was theorized to result from delayed exposure to early life infections in first born compared to later born children^26^. Importantly, assessment of birth order in prior contexts represents differences in leukemia risk across separate pregnancies, and thus are subject to variations in gestational influences across broad time periods. Twin plurality, in contrast, measures a shared gestation. Here, we find that DNA methylation markers associated with birth order^18^ differ significantly between first and second born twins, specifically in instances when an ALL case is born first. This suggests whatever biological mechanism leading to an increase in ALL risk in first born children may have similarities to that of first-born twins and given its reflection in DNA methylation patterns at birth may result from intrauterine or birth process influences. Interestingly the birth order direction of risk in AML is inverse to ALL, which also mirrors the inverse associations in singleton births^28^. Further investigation in a broader cohort of twins with acute leukemia is warranted.

The most important limitation to this study is the lack of zygosity information available in the evaluated registry data. This prevents accurate estimations of heritable genetic versus shared environmental influences on pediatric leukemia development. However, our estimation of monozygotic twin pairs allows for broad comparison to previously identified leukemia concordance rates. In a subset of twins with ALL from this cohort investigated using a DNA single nucleotide polymorphism array, approximately 50% of same sex pairs were monozygotic, which is consistent with the estimated results displayed here. In addition, given the low rates of concordant twin cancer diagnoses, there are multiple categories of cancer types for which we are underpowered to calculate SIR values. Furthermore, we are limited by the lack of recent registry data, raising potential for lead-time bias in twin siblings born in the latest birth years assessed. For instance, twins born in 2017 would only have cancer registry data available for 4 total years (through 2021), while latency in development of concordant leukemias has been shown to occur up to 9 years after the initial diagnosis in one sibling^25^. However, the inclusion of 33 years of cancer registry data and 35 years of birth registry data limits the impact of lead-time bias over the majority of our study period.

In conclusion, we identified twin childhood and AYA leukemia concordance rates through assessment of 40 years of California-based registry data. Concordance rates for acute leukemia were substantially lower than previous investigations, which may be a consequence of the unbiased, population-based approach utilized in this study or differences in the underlying risks of the ancestral populations composing the California cohort which is predominantly Latino. We further identified a unique association between first born twins and increased risk of childhood leukemia development, which was most notable for childhood ALL. This may relate to previous data suggesting a relationship between ALL, AML and birth order, however additional studies are necessary to determine the biological foundation of this result.

## Data Availability

The data that support the findings of this study are available from California Department of Public Health. Restrictions apply to the availability of these data, which were used under license for this study. Data are available from the authors with the permission of the California Department of Public Health. All available processed data are contained in the manuscript and its supplementary materials.

## ACKNOLWEDEMENTS

Funding provided by the V Foundation (FP067172) and the USC Norris Comprehensive Cancer Center SPORE development award (JLW).

## AUTHOR CONTRIBUTIONS

EMN: Data curation, methodology, statistical analysis, data visualization and manuscript writing. NZ: Statistical analysis, manuscript writing JLW: Methodology, manuscript writing

## CONFLICT OF INTEREST

The authors have no conflicts of interest to disclose.

## DATA AVAILABILITY

The data that support the findings of this study are available from California Department of Public Health. Restrictions apply to the availability of these data, which were used under license for this study. Data are available from the authors with the permission of the California Department of Public Health.

## ETHICS STATEMENT

The collection of cancer incidence data used in this study was supported by the California Department of Public Health as part of the statewide cancer reporting program mandated by California Health and Safety Code Section 103885; the National Cancer Institute’s Surveillance, Epidemiology, and End Results Program under contract HHSN261201000140C awarded to the Cancer Prevention Institute of California, contract HHSN261201000035C awarded to the University of Southern California, and contract HHSN261201000034C awarded to the Public Health Institute; and the Centers for Disease Control and Prevention’s National Program of Cancer Registries, under agreement U58DP003862-01 awarded to the California Department of Public Health. The ideas and opinions expressed herein are those of the author(s), and no endorsement by the California Department of Public Health, the National Cancer Institute, or the Centers for Disease Control and Prevention or their contractors and subcontractors is intended or should be inferred.

## Notes

### Competing Interest Statement

The authors have declared no competing interest.

### Author Declarations

This study was approved by Institutional Review Boards at the California Health and Human Services Agency Committee for the Protection of Human Subjects and the University of Southern California.

