## Supplemental Methods for "First-born twin has a higher risk of acute leukemia in a population-based assessment of cancer in twins in California, and lower than anticipated rate of twin concordance"

### *Standardized Incidence Ratio (SIR)*

Briefly, the SIR represents the relative risk of developing a cancer in healthy twin siblings of probands affected by a childhood or AYA cancer. SIRs were calculated in two manners. First, we assessed SIRs for any type of pediatric/AYA cancer development irrespective of the probands cancer diagnosis. We separately assessed SIRs for developing the same cancer type in healthy twin siblings as the proband. The first child to develop cancer in the twin pairs were defined as probands, and the standardized incidence ratios (SIRs) of cancer for the other child in the twin pairs (defined as "siblings") are calculated. This ratio is obtained by dividing the observed number of siblings with cancer by the expected number of cancers among siblings. To compute the expected number of siblings with cancer, the total years at risk are multiplied by the corresponding age group (5-year intervals) and sex-specific incidence rate of cancer, calculated individually and summed. Years at risk are individually determined by subtracting the age at the start of risk from the age at the end of risk. The age at the start of risk is defined as the year of cancer diagnosis in the probands. The age at the end of risk is defined was determined as either the end year of follow-up (2021) if the sibling remained cancer-free until the end of follow-up, or the year when the sibling received the cancer diagnosis. Age group and sex-specific incidence rates of cancer were obtained from SEER*Stat version 8.4.3, utilizing California data from SEER 17 Registries. The 95% confidence intervals (CIs) were computed assuming a Poisson distribution.

### *Birth order DNA methylation analysis*

We investigated a subset of monzygotic twin pairs discordant for acute lymphoblastic leukemia (n = 41) who previously underwent DNA methylation profiling using the Illumina EPIC BeadChip Array[17] from archived neonatal blood spots (i.e., prior to leukemia diagnosis. For regression modeling, beta values were $\log_2$ transformed to M-values. Significance was set at a false discovery rate (FDR) <0.05 to account for multiple comparisons. Linear regression modeling was conducted using the following equation:

$$DNA\ methylation\ Mvalue \sim Twin\ plurality\ order + ALL\ case\ status + sex + birth\ weight + twin\ pair\ ID + plate\ (batch\ effect) + nucleated\ cell\ proportions$$

To test for associations between twin plurality order and previously identified sites of DNA methylation associated with singleton birth order[18], we identified birth order CpGs that overlapped the EPIC array results from the discordant-ALL cohort. Beta values were used from the EPIC array, which correspond to the degree of DNA methylation at a particular CpG ranging from 0 (unmethylated) to 1 (fully methylated). Delta beta values were calculated as the first-born beta value minus the second-born beta value for each CpG site. Statistical comparisons between groups were made using a Wilcoxon rank-sum test.
