## Supplemental Tables for "First-born twin has a higher risk of acute leukemia in a population-based assessment of cancer in twins in California, and lower than anticipated rate of twin concordance"

**Table S1:** 12 broad groups and 69 subgroups classification per the International Classification of Childhood Cancer, Third Edition. Abbreviations used in manuscript are shown.

| Abbreviation | ICCC Category |
|---|---|
| **Leukemia** | **I. Leukemias, myeloproliferative diseases, and myelodysplastic diseases** |
| | *11. Lymphoid Leukemias* |
| | *12. Acute Myeloid Leukemias* |
| | *13. Chronic myeloproliferative diseases* |
| | *14. Myelodysplastic syndrome and Myeloproliferative diseases* |
| | *15. Unspecified and other specified leukemias* |
| **Lymphoma** | **II. Lymphomas and reticuloendothelial neoplasms** |
| | *21. Hodgkin lymphomas* |
| | *22. Non-Hodgkin lymphomas (except Burkitt lymphoma)* |
| | *23. Burkitt lymphoma* |
| | *24. Miscellaneous lymphoreticular neoplasms* |
| | *25. Unspecified lymphomas* |
| **CNS Neoplasms** | **III. CNS and miscellaneous intracranial and intraspinal neoplasms** |
| | *31. Ependymomas and choroid plexus tumor* |
| | *32. Astrocytomas* |
| | *33. Intracranial and intraspinal embryonal tumors* |
| | *34. Other Gliomas* |
| | *35. Other specified intracranial and intraspinal neoplasms* |
| | *36. Unspecified intracranial and intraspinal neoplasms* |
| **Neuroblastoma** | **IV. Neuroblastoma and other peripheral nervous cell tumors** |
| | *41. Neuroblastoma and ganglioneuroblastoma* |
| | *42. Other peripheral nervous cell tumors* |
| **Retinoblastoma** | **V. Retinoblastoma** |
| | *50. Retinoblastoma* |
| **Renal Tumors** | **VI. Renal Tumors** |
| | *61. Nephroblastoma and other nonepithelial renal tumors* |
| | *62. Renal carcinomas* |
| | *63. Unspecified malignant renal tumors* |
| **Hepatic Tumors** | **VII. Hepatic Tumors** |
| | *71. Hepatoblastoma* |
| | *72. Hepatic carcinomas* |
| | *73. Unspecified malignant hepatic tumors* |
| **Malignant Bone Tumors** | **VIII. Malignant bone tumors** |
| | *81. Osteosarcomas* |
| | *82. Chrondrosarcomas* |
| | *83. Ewing tumor and related sarcomas of bone* |
| | *84. Other specified malignant bone tumors* |
| | *85. Unspecified malignant bone tumors* |
| **Soft Tissue Sarcomas** | **IX. Soft tissue and other extraosseous sarcomas** |
| | *91. Rhabdomyosarcomas* |
| | *92. Fibrosarcomas, peripheral nerve sheath tumors, and other fibrous neoplasms* |
| | *93. Kaposi sarcomas* |
| | *94. Other specified soft tissue sarcomas* |
| | *95. Unspecified soft tissue sarcomas* |
| **Germ Cell Tumors** | **X. Germ cell tumors, trophoblastic tumors, and neoplasms of gonads** |
| | *101. Intracranial and intraspinal germ cell tumors* |
| | *102. Malignant extracranial and extragonadal germ cell tumors* |
| | *103. Malignant gonadal germ cell tumors* |
| | *104. Gonadal carcinomas* |
| | *105. Other and unspecified malignant gonadal tumors* |
| **Epithelial Neoplasms** | **XI. Other malignant epithelial neoplasms and malignant melanomas** |
| | *111. Adrenocortical carcinomas* |
| | *112. Thyroid carcinomas* |
| | *113. Nasopharyngeal carcinomas* |
| | *114. Malignant melanomas* |
| | *115. Skin carcinomas* |
| | *116. Other and unspecified carcinomas* |
| **Other Neoplasms** | **XII. Other and unspecified malignant neoplasms** |
| | *121. Other specified malignant tumors* |
| | *122. Other unspecified malignant tumors* |

**Table S2:** Mean birthweight values and twin birth order by pediatric cancer case status. Data from concordant pairs is not included in table. P-values for birth weight result from paired T-test. Odds ratios and P-values for birth order result from Fishers exact test. SD = standard deviation. OR = odds ratio. * = P<0.05, **=P<0.01.

| | Birthweight (SD) | | | Case Birth Order | | | |
|---|---|---|---|---|---|---|---|
| **Category** | ***Cases*** | ***Controls*** | ***P*** | ***First*** | ***Second*** | ***OR (95% CI)*** | ***P*** |
| **All cancers** | 2448 (603) | 2438 (604) | 0.840 | 599 | 558 | 1.18 (1-1.39) | 0.049* |
| **By cancer group** | | | | | | | |
| Leukemias | 2501 (514) | 2473 (524) | 1 | 144 | 110 | 1.73 (1.20-2.52) | 0.003** |
| Lymphomas | 2561 (574) | 2553 (591) | 0.603 | 75 | 57 | 1.70 (1.02-2.87) | 0.036* |
| CNS Neoplasms | 2427 (623) | 2412 (578) | 0.791 | 106 | 123 | 0.70 (0.47-1.03) | 0.072 |
| Neuroblastoma | 2405 (558) | 2381 (618) | 0.678 | 25 | 29 | 0.84 (0.32-1.70) | 0.557 |
| Retinoblastoma | 2529 (651) | 2414 (589) | 0.607 | 10 | 9 | 1.27 (0.26-6.42) | 1 |
| Renal Tumors | 2404 (599) | 2354 (583) | 1 | 24 | 15 | 2.53 (0.94-7.05) | 0.069 |
| Hepatic Tumors | 1488 (824) | 1474 (841) | 1 | 18 | 15 | 1.43 (0.49-4.24) | 0.622 |
| Malignant bone tumors | 2477 (544) | 2480 (498) | 0.401 | 31 | 19 | 2.64 (1.10-6.47) | 0.027* |
| Soft Tissue Sarcomas | 2490 (530) | 2467 (497) | 0.391 | 41 | 50 | 0.66 (0.35-1.25) | 0.225 |
| Germ Cell Tumors | 2419 (618) | 2444 (585) | 1 | 47 | 42 | 1.25 (0.67-2.35) | 0.549 |
| Epithelial Neoplasms | 2482 (583) | 2508 (637) | 0.629 | 101 | 105 | 0.93 (0.62-1.39) | 0.768 |
| Other Neoplasms | 2589 (559) | 2503 (562) | 0.388 | 9 | 5 | 3.76 (0.56-30.40) | 0.220 |

**Table S3:** Twin pediatric cancer concordance and discordance for all cancers and by ICCC-3 broad groups and subgroups diagnostic groups and subgroups. Data for all subgroups, including those without concordant twin pairs, is shown. Con = Concordant. Dis = Discordant. % = Percent Concordance.

| | | Same Sex | | | Opposite Sex | | | All | | |
|---|---|---|---|---|---|---|---|---|---|---|
| | **N (pairs)** | **Con** | **Dis** | **%** | **Con** | **Dis** | **%** | **Con** | **Dis** | **%** |
| **All cancers** | **1213** | **10** | **826** | **1.2** | **3** | **374** | **0.8** | **13** | **1200** | **1.1** |
| **Leukemias** | **255** | **2** | **164** | **1.2** | **0** | **89** | **0** | **2** | **253** | **0.8** |
| *11. Lymphoid Leukemias* | 199 | 1 | 133 | 0.8 | 0 | 65 | 0 | 1 | 198 | 0.5 |
| *12. Acute Myeloid Leukemias* | 34 | 1 | 19 | 5.0 | 0 | 14 | 0 | 1 | 33 | 3.0 |
| *13. Chronic myeloproliferative diseases* | 11 | 0 | 8 | 0 | 0 | 3 | 0 | 0 | 11 | 0 |
| *14. Myelodysplastic syndrome and myeloproliferative diseases* | 8 | 0 | 3 | 0 | 0 | 5 | 0 | 0 | 8 | 0 |
| *15. Unspecified and other specified leukemias* | 4 | 0 | 1 | 0 | 0 | 3 | 0 | 0 | 4 | 0 |
| **Lymphomas** | **134** | **0** | **89** | **0** | **1** | **44** | **2.2** | **1** | **133** | **0.7** |
| *21. Hodgkin lymphomas* | 71 | 0 | 47 | 0 | 0 | 24 | 0 | 0 | 71 | 0 |
| *22. Non-Hodgkin lymphomas (except Burkitt lymphoma)* | 53 | 0 | 33 | 0 | 1 | 19 | 5.0 | 1 | 52 | 1.9 |
| *23. Burkitt lymphoma* | 12 | 0 | 10 | 0 | 0 | 2 | 0 | 0 | 12 | 0 |
| *24. Miscellaneous lymphoreticular neoplasms* | 0 | 0 | 0 | NA | 0 | 0 | NA | 0 | 0 | NA |
| *25. Unspecified lymphomas* | 0 | 0 | 0 | NA | 0 | 0 | NA | 0 | 0 | NA |
| **CNS Neoplasms** | **232** | **1** | **162** | **0.6** | **0** | **69** | **0.0** | **1** | **231** | **0.4** |
| *31. Ependymomas and choroid plexus tumor* | 16 | 0 | 12 | 0 | 0 | 4 | 0 | 0 | 16 | 0 |
| *32. Astrocytomas* | 88 | 0 | 60 | 0 | 0 | 28 | 0 | 0 | 88 | 0 |
| *33. Intracranial and intraspinal embryonal tumors* | 15 | 0 | 12 | 0 | 0 | 3 | 0 | 0 | 15 | 0 |
| *34. Other Gliomas* | 26 | 0 | 15 | 0 | 0 | 11 | 0 | 0 | 26 | 0 |
| *35. Other specified intracranial and intraspinal neoplasms* | 72 | 1 | 48 | 2.0 | 0 | 23 | 0 | 1 | 71 | 1.4 |
| *36. Unspecified intracranial and intraspinal neoplasms* | 5 | 0 | 3 | 0 | 0 | 2 | 0 | 0 | 5 | 0 |
| **Neuroblastoma** | **54** | **1** | **41** | **2.4** | **0** | **12** | **0** | **1** | **53** | **1.9** |
| *41. Neuroblastoma and ganglioneuroblastoma* | 52 | 1 | 39 | 2.5 | 0 | 12 | 0 | 1 | 51 | 2.0 |
| *42. Other peripheral nervous cell tumors* | 2 | 0 | 2 | 0 | 0 | 0 | NA | 0 | 2 | 0 |
| **Retinoblastoma** | **17** | **1** | **11** | **8.3** | **1** | **4** | **20.0** | **2** | **15** | **11.8** |
| *50. Retinoblastoma* | 16 | 1 | 11 | 8.3 | 1 | 3 | 25.0 | 2 | 14 | 12.5 |
| **Renal Tumors** | **39** | **0** | **21** | **0** | **0** | **18** | **0** | **0** | **39** | **0** |
| *61. Nephroblastoma and other nonepithelial renal tumors* | 39 | 0 | 21 | 0 | 0 | 18 | 0 | 0 | 39 | 0 |
| *62. Renal carcinomas* | 0 | 0 | 0 | NA | 0 | 0 | NA | 0 | 0 | NA |
| *63. Unspecified malignant renal tumors* | 0 | 0 | 0 | NA | 0 | 0 | NA | 0 | 0 | NA |
| **Hepatic Tumors** | **33** | **0** | **23** | **0** | **0** | **10** | **0** | **0** | **33** | **0** |
| *71. Hepatoblastoma* | 29 | 0 | 20 | 0 | 0 | 9 | 0 | 0 | 29 | 0 |
| *72. Hepatic carcinomas* | 4 | 0 | 3 | 0 | 0 | 1 | 0 | 0 | 4 | 0 |
| *73. Unspecified malignant hepatic tumors* | 0 | 0 | 0 | NA | 0 | 0 | NA | 0 | 0 | NA |
| **Malignant bone tumors** | **54** | **0** | **36** | **0** | **0** | **18** | **0** | **0** | **54** | **0** |
| *81. Osteosarcomas* | 32 | 0 | 21 | 0 | 0 | 11 | 0 | 0 | 32 | 0 |
| *82. Chrondrosarcomas* | 2 | 0 | 2 | 0 | 0 | 0 | NA | 0 | 2 | 0 |
| *83. Ewing tumor and related sarcomas of bone* | 19 | 0 | 13 | 0 | 0 | 6 | 0 | 0 | 19 | 0 |
| *84. Other specified malignant bone tumors* | 1 | 0 | 0 | NA | 0 | 1 | 0 | 0 | 1 | 0 |
| *85. Unspecified malignant bone tumors* | 0 | 0 | 0 | NA | 0 | 0 | NA | 0 | 0 | NA |
| **Soft Tissue Sarcomas** | **92** | **1** | **67** | **1.5** | **0** | **24** | **0** | **1** | **91** | **1.1** |
| *91. Rhabdomyosarcomas* | 29 | 0 | 21 | 0 | 0 | 8 | 0 | 0 | 29 | 0 |
| *92. Fibrosarcomas, peripheral nerve sheath tumors, and other fibrous neoplasms* | 14 | 0 | 10 | 0 | 0 | 4 | 0 | 0 | 14 | 0 |
| *93. Kaposi sarcomas* | 1 | 0 | 1 | 0 | 0 | 0 | NA | 0 | 1 | 0 |
| *94. Other specified soft tissue sarcomas* | 41 | 1 | 29 | 3.3 | 0 | 11 | 0 | 1 | 40 | 2.4 |
| *95. Unspecified soft tissue sarcomas* | 7 | 0 | 6 | 0 | 0 | 1 | 0 | 0 | 7 | 0 |
| **Germ Cell Tumors** | **95** | **3** | **73** | **3.9** | **0** | **19** | **0** | **3** | **92** | **3.2** |
| *101. Intracranial and intraspinal germ cell tumors* | 12 | 0 | 8 | 0 | 0 | 4 | 0 | 0 | 10 | 0 |
| *102. Malignant extracranial and extragonadal germ cell tumors* | 6 | 0 | 6 | 0 | 0 | 0 | NA | 0 | 6 | 0 |
| *103. Malignant gonadal germ cell tumors* | 67 | 3 | 51 | 5.6 | 0 | 13 | 0 | 3 | 67 | 4.3 |
| *104. Gonadal carcinomas* | 8 | 0 | 7 | 0 | 0 | 1 | 0 | 0 | 8 | 0 |
| *105. Other and unspecified malignant gonadal tumors* | 2 | 0 | 1 | 0 | 0 | 1 | 0 | 0 | 2 | 0 |
| **Epithelial Neoplasms** | **214** | **1** | **143** | **0.7** | **1** | **69** | **1.4** | **2** | **212** | **0.9** |
| *111. Adrenocortical carcinomas* | 1 | 0 | 0 | NA | 0 | 1 | 0 | 0 | 1 | 0 |
| *112. Thyroid carcinomas* | 74 | 1 | 48 | 2.1 | 1 | 24 | 4.0 | 2 | 72 | 2.8 |
| *113. Nasopharyngeal carcinomas* | 1 | 0 | 2 | 0 | 0 | 0 | NA | 0 | 2 | 0 |
| *114. Malignant melanomas* | 56 | 0 | 38 | 0 | 0 | 18 | 0 | 0 | 56 | 0 |
| *115. Skin carcinomas* | 6 | 0 | 4 | 0 | 0 | 2 | 0 | 0 | 6 | 0 |
| *116. Other and unspecified carcinomas* | 87 | 0 | 61 | 0 | 0 | 26 | 0 | 0 | 87 | 0 |
| **Other Neoplasms** | **13** | **0** | **9** | **0** | **0** | **4** | **0** | **0** | **13** | **0** |
| *121. Other specified malignant tumors* | 5 | 0 | 3 | 0 | 0 | 2 | 0 | 0 | 5 | 0 |
| *122. Other unspecified malignant tumors* | 8 | 0 | 6 | 0 | 0 | 2 | 0 | 0 | 8 | 0 |

**Table S4:** Twin birth order by pediatric leukemia case status across recode subgroups. Odds ratios represent odds of the case being first born in a twin pair. P-values result from Fishers exact test. OR = odds ratio. * = P<0.05, ***=P<0.001.

| Category | OR (95% CI) | P |
|---|---|---|
| **Leukemias** | | |
| *Lymphoid Leukemias* | 2.21 (1.44-3.39) | 0.000165*** |
| *Acute myeloid Leukemias* | 0.41 (0.13-1.24) | 0.127 |
| *Chronic myeloproliferative diseases* | 6.41 (0.82-69.21) | 0.086 |
| *Myelodysplastic syndrome and other myeloproliferative diseases* | 1 (0.13-10.44) | 1 |
| *Unspecified and other specific leukemias* | 0.16 (0.001-4.72) | 0.486 |

**Table S5:** Estimation of twin cancer concordance rates by zygosity status by ICCC-3 broad groups and subgroups diagnostic groups and subgroups. Data for all subgroups, including those without concordant twin pairs, is shown. Con = Concordant. Dis = Discordant. % = Percent Concordance.

| | **Observed Data (N)** | | | **Estimated Monozygotic** | | | | **Estimated Dizygotic** | | | |
|---|---|---|---|---|---|---|---|---|---|---|---|
| | *Total* | *Same Sex* | *Opposite Sex* | *N* | *Con* | *Dis* | *%* | *N* | *Con* | *Dis* | *%* |
| **All cancers** | 1213 | 836 | 377 | 460 | 10 | 450 | 2.2 | 753 | 3 | 750 | 0.4 |
| **By cancer group** | | | | | | | | | | | |
| **Leukemias** | **255** | **166** | **89** | **77** | **2** | **75** | **2.6** | **178** | **0** | **180** | **0.0** |
| *11. Lymphoid Leukemias* | 199 | 134 | 65 | 69 | 1 | 68 | 1.4 | 130 | 0 | 130 | 0.0 |
| *12. Acute Myeloid Leukemias* | 34 | 20 | 14 | 6 | 1 | 5 | 16.7 | 28 | 0 | 28 | 0.0 |
| **Lymphomas** | **134** | **89** | **45** | **44** | **0** | **44** | **0.0** | **90** | **1** | **89** | **1.1** |
| *22. Non-Hodgkin lymphomas (except Burkitt lymphoma)* | 53 | 33 | 20 | 13 | 0 | 33 | 0.0 | 40 | 1 | 39 | 2.5 |
| **CNS Neoplasms** | **232** | **163** | **69** | **94** | **1** | **93** | **1.1** | **138** | **0** | **138** | **0.0** |
| *35. Other specified intracranial and intraspinal neoplasms* | 72 | 49 | 23 | 26 | 1 | 25 | 4.0 | 46 | 0 | 46 | 0.0 |
| **Neuroblastoma** | **54** | **42** | **12** | **30** | **1** | **29** | **3.3** | **24** | **0** | **24** | **0.0** |
| *41. Neuroblastoma and ganglioneuroblastoma* | 52 | 40 | 12 | 28 | 1 | 27 | 3.6 | 24 | 0 | 24 | 0.0 |
| **Retinoblastoma** | **17** | **12** | **5** | **7** | **1** | **6** | **14.3** | **10** | **1** | **9** | **10.0** |
| *50. Retinoblastoma* | 17 | 12 | 5 | 7 | 1 | 6 | 14.3 | 10 | 1 | 9 | 10.0 |
| **Renal Tumors** | **39** | **21** | **18** | **3** | **0** | **3** | **0.0** | **36** | **0** | **18** | **0.0** |
| **Hepatic Tumors** | **33** | **23** | **10** | **13** | **0** | **13** | **0.0** | **10** | **0** | **10** | **0.0** |
| **Malignant bone tumors** | **54** | **36** | **18** | **18** | **0** | **18** | **0.0** | **18** | **0** | **18** | **0.0** |
| **Soft Tissue Sarcomas** | **92** | **68** | **24** | **44** | **1** | **43** | **2.3** | **48** | **0** | **48** | **0.0** |
| *94. Other specified soft tissue sarcomas* | 41 | 30 | 11 | 19 | 1 | 18 | 5.3 | 22 | 0 | 22 | 0.0 |
| **Germ Cell Tumors** | **95** | **75** | **19** | **56** | **3** | **53** | **5.4** | **39** | **0** | **39** | **0.0** |
| *103. Malignant gonadal germ cell tumors* | 67 | 54 | 13 | 41 | 3 | 38 | 7.3 | 26 | 0 | 26 | 0.0 |
| **Epithelial Neoplasms** | **214** | **144** | **70** | **74** | **1** | **73** | **1.4** | **140** | **1** | **139** | **0.7** |
| *112. Thyroid carcinomas* | 74 | 49 | 25 | 24 | 1 | 23 | 4.2 | 50 | 1 | 49 | 2.0 |
| **Other Neoplasms** | **13** | **9** | **4** | **5** | **0** | **9** | **0.0** | **8** | **0** | **4** | **0.0** |

**Table S6:** Standardized incidence ratios (SIRs) for cancer in twins showing risk of development of the same cancer type (by recode subgroup) in the twin sibling of probands. CI=confidence interval.

| Category | Probands | Observed | Expected | SIR (95% CI) |
|---|---|---|---|---|
| **Broad groups** | | | | |
| Leukemias | 254 | 2 | 0.13 | 15.6 (1.9 - 56.4) |
| Lymphomas | 134 | 1 | 0.06 | 16.3 (0.4 - 90.7) |
| CNS Neoplasms | 233 | 1 | 0.06 | 17.2 (0.4 - 95.6) |
| Neuroblastoma | 54 | 1 | 0.004 | 249.9 (6.3 - 1392.4) |
| Retinoblastoma | 17 | 2 | 0.0006 | 3166.0 (383.4 - 11436.9) |
| Renal Tumors | 39 | 0 | | |
| Hepatic Tumors | 33 | 0 | | |
| Malignant bone tumors | 54 | 0 | | |
| Soft Tissue Sarcomas | 91 | 1 | 0.02 | 62.4 (1.6 - 347.9) |
| Germ Cell Tumors | 95 | 3 | 0.04 | 66.7 (13.8 - 195) |
| Epithelial Neoplasms | 214 | 2 | 0.37 | 5.4 (0.7 - 19.7) |
| Other Neoplasms | 7 | 0 | | |

**Table S7:** Standardized incidence ratios (SIRs) of risk of any cancer development in siblings of twins with cancer for any pediatric or adolescent/young adult cancer type. Results are displayed by broad cancer group diagnosed in the proband, and SIRs are based on risk of any cancer type in the proband.

| | Probands | Observed | Expected | SIR (95% CI) |
|---|---|---|---|---|
| **All Cancer** | 1213 | 15 | 3.47 | 4.3 (2.4 - 7.1) |
| **Broad groups** | | | | |
| Leukemias | 254 | 2 | 0.69 | 2.9 (0.4, 10.5) |
| Lymphomas | 134 | 1 | 0.41 | 2.5 (0.1, 13.7) |
| CNS Neoplasms | 233 | 1 | 0.62 | 1.6 (0.0, 9.0) |
| Neuroblastoma | 54 | 1 | 0.14 | 7.2 (0.2, 40.0) |
| Retinoblastoma | 17 | 2 | 0.07 | 29.2 (3.5, 105.4) |
| Renal Tumors | 39 | 0 | | |
| Hepatic Tumors | 33 | 0 | | |
| Malignant bone tumors | 54 | 1 | 0.16 | 6.4 (0.2, 35.4) |
| Soft Tissue Sarcomas | 91 | 2 | 0.29 | 6.8 (0.8, 24.6) |
| Germ Cell Tumors | 95 | 4 | 0.26 | 15.5 (4.2, 39.7) |
| Epithelial Neoplasms | 214 | 2 | 0.67 | 3.0 (0.4, 10.8) |
| Other Neoplasms | 12 | 0 | | |

**Table S8:** Cell type composition analysis by twin birth order. A referenced deconvolution analysis (IDOL) was used to determine the proportions of nucleated cells from archived neonatal dried blood spot samples using DNA methylation profiling from 41 twin pairs discordant for pediatric acute lymphoblastic leukemia. Paired T-test was used to evaluate for differences in cell proportion for each nucleated cell type assessed. SD = Standard deviation.

| Cell type | First born mean (SD) | Second born mean (SD) | P |
|---|---|---|---|
| B cells | 0.050 (0.022) | 0.049 (0.025) | 0.815 |
| CD4 T-cells | 0.209 (0.070) | 0.212 (0.089) | 0.623 |
| CD8 T-cells | 0.041 (0.026) | 0.040 (0.028) | 0.804 |
| NK cells | 0.009 (0.013) | 0.011 (0.016) | 0.175 |
| Monocytes | 0.087 (0.038) | 0.087 (0.031) | 0.905 |
| Granulocytes | 0.558 (0.118) | 0.553 (0.141) | 0.682 |
| Nucleated red blood cells | 0.036 (0.120) | 0.037 (0.139) | 0.884 |

**Table S9:** Top 20 significant differentially methylated probes at birth identified from birth plurality order linear regression analysis in 41 ALL-discordant twin pairs. Positions referenced to Hg19.

| CpG | Estimate | Std. Error | Pr(>\|t\|) | FDR | Chromosome | Position | UCSC_RefGene_Name |
|---|---|---|---|---|---|---|---|
| cg26335251 | -45.606557 | 5.89765682 | 1.41E-07 | 0.02548192 | chr17 | 75539921 | NA |
| cg10817023 | -21.832225 | 2.89403871 | 2.08E-07 | 0.02548192 | chr5 | 133853517 | NA |
| cg19442702 | -22.309636 | 2.95434736 | 2.05E-07 | 0.02548192 | chr5 | 398425 | AHRR |
| cg01321488 | -21.106998 | 2.74806767 | 1.57E-07 | 0.02548192 | chr9 | 127380298 | NR6A1;NR6A1;NR6A1 |
| cg09738193 | -21.973898 | 2.94816072 | 2.51E-07 | 0.02548192 | chr16 | 67926317 | PSKH1 |
| cg26618645 | -34.533609 | 4.63063711 | 2.49E-07 | 0.02548192 | chr16 | 30780332 | RNF40 |
| cg01813254 | -27.728941 | 3.49704088 | 9.49E-08 | 0.02548192 | chr11 | 9036548 | NA |
| cg19262334 | -29.220747 | 3.9599124 | 2.93E-07 | 0.02602671 | chr18 | 74631861 | ZNF236 |
| cg05573381 | -30.678108 | 4.23816372 | 3.94E-07 | 0.02796016 | chr5 | 1653258 | NA |
| cg05052194 | -29.266438 | 4.03893695 | 3.88E-07 | 0.02796016 | chr1 | 2160249 | SKI |
| cg18690833 | -34.167392 | 4.87896345 | 6.49E-07 | 0.02949527 | chr20 | 13972022 | SEL1L2 |
| cg10329345 | -22.583062 | 3.18165365 | 5.30E-07 | 0.02949527 | chr1 | 45083079 | RNF220 |
| cg09407429 | -22.558022 | 3.21493351 | 6.30E-07 | 0.02949527 | chr3 | 4534383 | ITPR1;ITPR1;ITPR1 |
| cg25528260 | -38.485094 | 5.50421806 | 6.65E-07 | 0.02949527 | chr7 | 72777727 | NA |
| cg07876051 | -38.942799 | 5.53220661 | 6.01E-07 | 0.02949527 | chr10 | 3159145 | PFKP |
| cg07805959 | -18.607996 | 2.62367993 | 5.37E-07 | 0.02949527 | chr17 | 2595004 | KIAA0664 |
| cg27120649 | -31.710823 | 4.59687307 | 8.12E-07 | 0.03393192 | chr7 | 94286261 | SGCE;PEG10;SGCE;PEG10;SGCE |
| cg07607077 | -19.43779 | 3.30165743 | 7.63E-06 | 0.03408781 | chr7 | 919584 | C7orf20 |
| cg04376617 | 32.6600413 | 5.25666516 | 3.66E-06 | 0.03408781 | chr18 | 43685360 | ATP5A1;HAUS1;HAUS1 |
| cg12285834 | -31.06016 | 4.7980595 | 2.05E-06 | 0.03408781 | chr3 | 72345017 | NA |

**Table S10:** Top 20 significant differentially methylated regions at birth identified from Comb P analysis of birth plurality order linear regression analysis in 41 ALL-discordant twin pairs. Positions referenced to Hg19.

| Chromosome | Start | End | min_p | n_probes | z_p | z_sidak_p | Gene |
|---|---|---|---|---|---|---|---|
| chr16 | 89349945 | 89352007 | 0.007374 | 10 | 1.30E-10 | 4.46E-08 | *ANKRD11* |
| chr16 | 89437341 | 89439497 | 0.009317 | 12 | 1.46E-09 | 4.81E-07 | *ANKRD11* |
| chr5 | 169658215 | 169659950 | 0.001232 | 16 | 1.20E-09 | 4.92E-07 | NA |
| chr19 | 48674931 | 48676053 | 0.001771 | 6 | 1.62E-09 | 1.02E-06 | *ZSWIM9* |
| chr2 | 168724333 | 168726607 | 0.01452 | 9 | 6.68E-09 | 2.09E-06 | *B3GALT1* |
| chr17 | 77951408 | 77952463 | 0.002708 | 7 | 3.60E-09 | 2.43E-06 | *TBC1D1**6*** |
| chr13 | 113718453 | 113719440 | 2.67E-04 | 5 | 3.57E-09 | 2.57E-06 | *MCF2L* |
| chr1 | 68516272 | 68517691 | 4.44E-04 | 15 | 7.42E-09 | 3.71E-06 | *DIRAS3* |
| chr2 | 228497892 | 228499386 | 0.004894 | 6 | 8.25E-09 | 3.92E-06 | *C2orf83* |
| chr17 | 74303724 | 74304023 | 9.65E-05 | 8 | 1.84E-09 | 4.38E-06 | *QRICH2* |
| chr12 | 131271323 | 131272183 | 0.001841 | 3 | 7.46E-09 | 6.16E-06 | NA |
| chr5 | 170883661 | 170884806 | 0.01135 | 6 | 9.94E-09 | 6.17E-06 | *FGF18* |
| chr19 | 46917727 | 46918365 | 0.005363 | 3 | 6.03E-09 | 6.71E-06 | NA |
| chr6 | 87804765 | 87806133 | 0.001279 | 8 | 1.37E-08 | 7.11E-06 | *CGA* |
| chr15 | 69221572 | 69223018 | 0.007071 | 8 | 1.58E-08 | 7.78E-06 | *MIR548H4* |
| chr1 | 192917420 | 192918151 | 0.002707 | 4 | 8.35E-09 | 8.11E-06 | NA |
| chr17 | 17124521 | 17124969 | 1.55E-04 | 6 | 6.23E-09 | 9.87E-06 | *PLD6; FLCN* |
| chr12 | 101752716 | 101754080 | 0.002954 | 5 | 2.17E-08 | 1.13E-05 | *UTP20* |
| chr11 | 118501494 | 118502454 | 0.002753 | 6 | 1.90E-08 | 1.40E-05 | *PHLDB1* |
| chr2 | 106959205 | 106959878 | 8.18E-04 | 7 | 1.35E-08 | 1.42E-05 | NA |
