## Supplemental Figures for "First-born twin has a higher risk of acute leukemia in a population-based assessment of cancer in twins in California, and lower than anticipated rate of twin concordance"

**Figure S1:** Deconvolution analysis of 41 twin pairs discordant for pediatric acute lymphoblastic leukemia by plurality order.

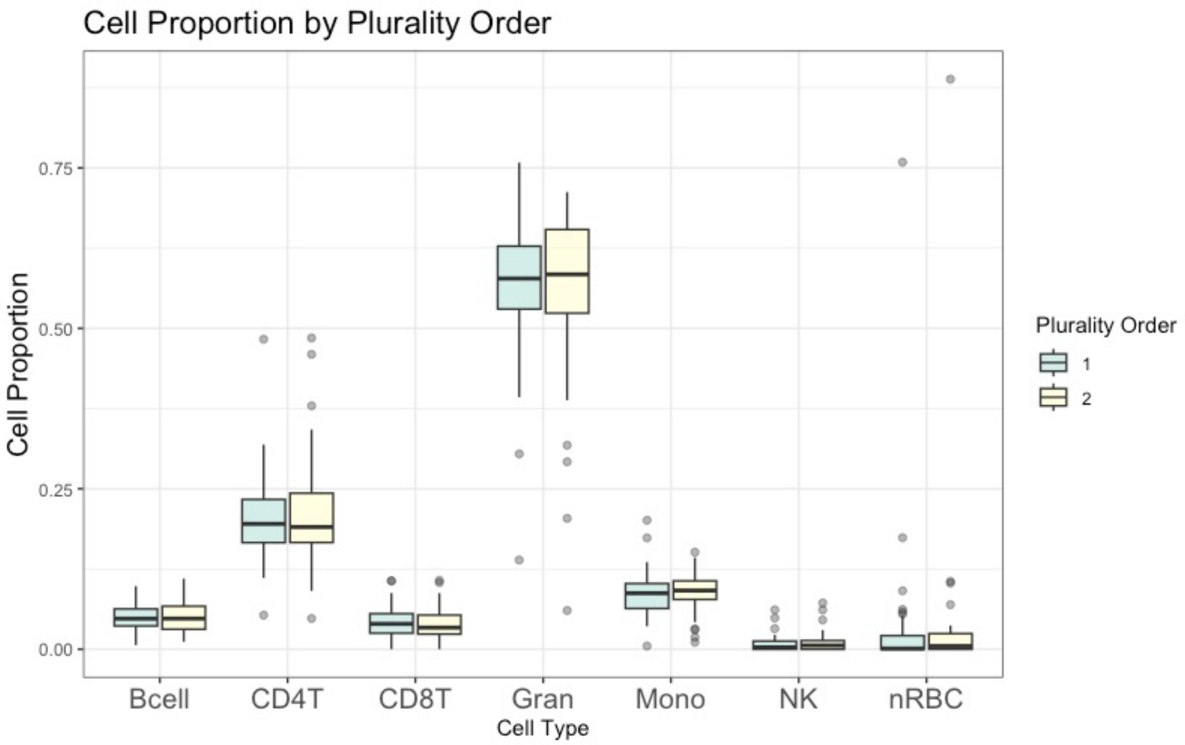

**Figure S2**: Distribution of plurality delta-beta (first-born beta minus second-born) for 305 birth order associated CpGs in 41 ALL-discordant twin pairs (82 individuals) assessed by the EPIC DNA methylation array. The plot is separated by instances in which cases are first-born (n = 24 twin pairs) and where cases are second-born (n = 17 twin pairs). Mean delta beta values are significantly different in twin sets in which the case is first born (n = 24) compared to those where the case is born second (n = 17, Wilcoxon rank sum P = 8.53e-12). Mean delta beta values were significantly lower than zero (hypomethylated) across the twin pairs in which the ALL case was first-born (P = $5.11 \times 10^{-8}$), while values were significantly higher than zero (hypermethylated) in pairs in which the case was second born (P = $5.04 \times 10^{-6}$).

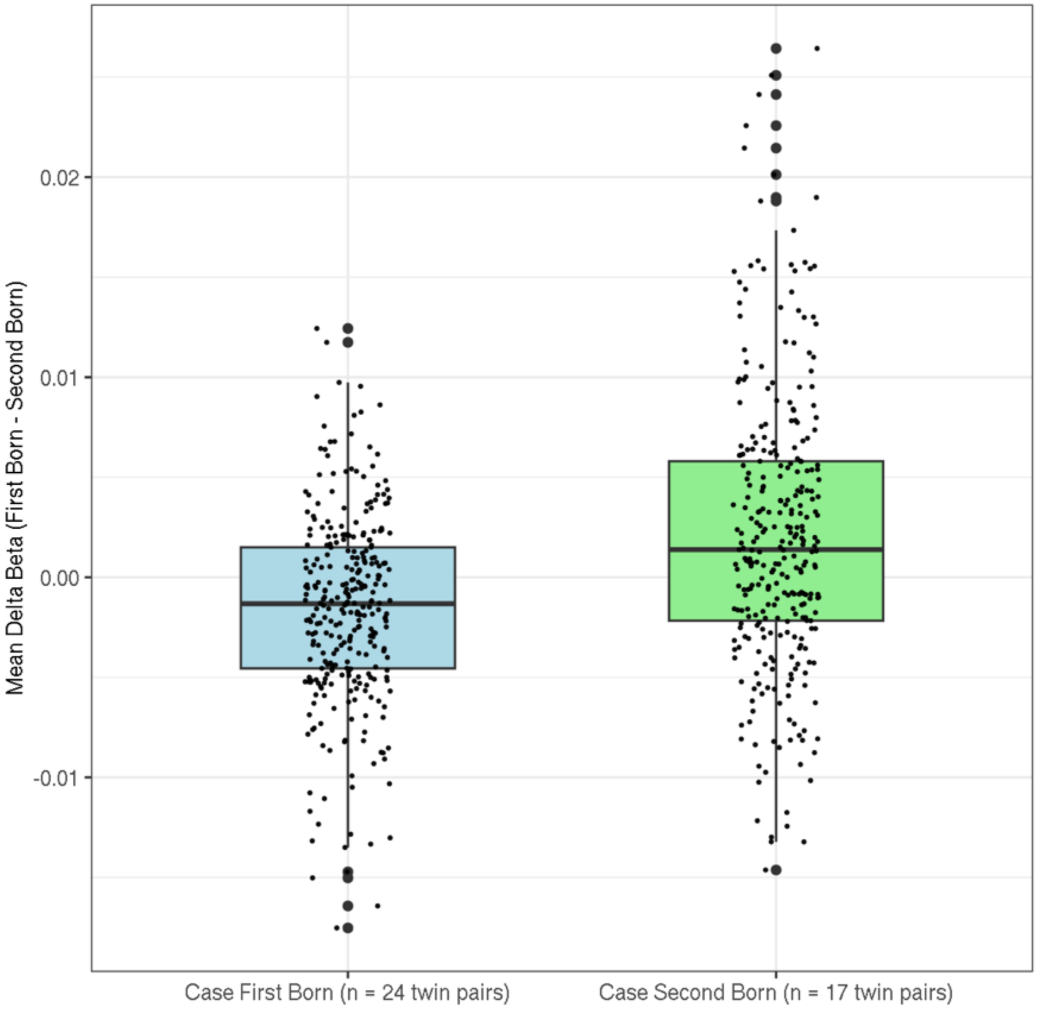
